# Pre-endoscopy SARS-CoV-2 testing strategy during COVID-19 pandemic: *The care must go on*

**DOI:** 10.1101/2020.10.22.20217885

**Authors:** M Casper, MC Reichert, J Rissland, S Smola, F Lammert, M Krawczyk

**Author notes:** **Correspondence address:** Markus Casper, MD, Saarland University Medical Center, Kirrberger Straße 100, D-66421 Homburg, Germany.

## Abstract

**Background:** In response to the COVID-19 pandemic, endoscopic societies have recommended reduction of endoscopic procedures. In particular non-urgent endoscopies should be postponed. However, this might lead to unnecessary delay in diagnosing gastrointestinal conditions.

**Methods:** Retrospectively we analysed the gastrointestinal endoscopies performed at the Central Endoscopy Unit of Saarland University Medical Center during seven weeks from 23 March to 10 May 2020 and present our real-world single-center experience with an individualized rtPCR-based pre-endoscopy SARS-CoV-2 testing (“PECo”) strategy.

**Results:** Altogether 359 gastrointestinal endoscopies were performed. The PECo strategy enabled us to conservatively handle endoscopy program reduction (44% reduction as compared 2019). The results of COVID-19 rtPCR from nasopharyngeal swabs were available in 89% of patients prior to endoscopies. Apart from six patients with known COVID-19, all other tested patients were negative. The frequencies of endoscopic therapies and clinically significant findings did not differ between patients with or without SARS-CoV-2 tests.

**Conclusion:** A reasonable reduction of the endoscopy program in the setting of structured SARS-CoV-2 testing is feasible and safe. The PECo strategy allows continuation of endoscopic procedures in a region with intermediate frequency of COVID-19 when hospital capacities are not overwhelmed by the pandemic. Thus, the study might help to develop new strategies during future waves of COVID-19 or local outbreaks.

## INTRODUCTION

COVID-19 substantially affects health care systems worldwide. Gastroenterologists must balance the risk for SARS-CoV-2 virus transmission during endoscopy against the risk caused by the delay of the procedures. Endoscopic societies around the world have published recommendations to assist endoscopists in decision-making during the pandemic (1). In most of these recommendations it is advised to use personal protective equipment (PPE) and to postpone non-urgent or routine procedures temporarily. However, urgent procedures are not uniformly defined, requiring case-by-case decisions from endoscopists in many patients. Following these recommendations, endoscopy programs were substantially reduced worldwide (on average by 83%) during the pandemic (2). The short- and medium-term consequences of this approach are difficult to estimate. As an alternative we present here our real-world experience with the rtPCR-based pre-endoscopy SARS-CoV-2 testing (“PECo”) strategy that enabled us to conservatively handle endoscopy program reduction.

## PATIENTS AND METHODS

### Study cohort and background

Retrospectively we analysed the gastrointestinal endoscopies performed at the Central Endoscopy Unit of Saarland University Medical Center during seven weeks from 23 March to 10 May 2020 (local peak period of the COVID-19 pandemic). This endoscopy unit is an academic tertiary referral centre serving all medical and surgical departments as well as state and regional health care providers. In Germany, more than 169.000 COVID-19 cases leading to more than 7.400 deaths (4.4%) had been reported until 10 May 2020. The current cumulative incidence in Saarland, which is one of the Federal states in Southwestern Germany at the border to France, is 279 cases per 100.000. Between 23 March and 10 May 2020, 2.514 individuals were tested positive for SARS-CoV-2 in Saarland, of whom 146 (5.8%) died.

### Testing strategy

The rtPCR-based pre-endoscopy SARS-CoV-2 testing (PECo) on nasopharyngeal swabs (Roche, Basel, Switzerlanld Altona Diagnostics, Hamburg, Germany) was broadly offered to in-patients in the Saarland University Medical Centre during the pandemic. Starting on 2 April 2020, structured rtPCR testing was also implemented for all out-patients scheduled for endoscopic procedures. Endoscopies were planned to be performed when the patient tested negative within 5 days prior to the procedure. In addition, all members of the endoscopy suite (nurses, physicians, endoscope reprocessing and cleaning staff) were tested weekly using pooling of samples, as described recently (3).

Routine rtPCR test results were available within 3 - 5 hours, when performed before 04:30 PM on weekdays or 02:00 PM on weekends, or on the next day when performed later. Emergency testing was available 24/7 within 3 hours. Patients were also inquired using questionnaires for typical COVID-19 symptoms prior to endoscopy. All necessary in-patient endoscopic procedures were performed as requested. For out-patients urgent procedures were unrestrictedly offered, non-urgent procedures were discussed on an individual basis with patients and referring physicians.

Risk-stratified personal protective measures conformed with the published recommendations (4). Asymptomatic patients with a negative rtPCR test result were categorized as low-risk patients and PPE was adapted (surgical mask, goggles, single-use gown, gloves, hairnet).

## RESULTS

During the analysed period, we performed 359 gastrointestinal endoscopic procedures. In the same period in 2019 a total of 626 (42.7% reduction) endoscopies were done (187 including 10 PEGs vs. 340 upper endoscopies [55%], 121 vs. 212 lower endoscopies [57.1%], 42 vs. 58 ERCPs [72.4%]). In total, 263 (73.2%) procedures were in-patient endoscopies (2019: 423; 67.6%). Six endoscopies performed in COVID-19 patients on extracorporeal membrane oxygenation were excluded from further analysis. Table 1 summarizes the indications, outcomes and baseline characteristics of 353 endoscopies included in the final analysis. The majority of endoscopies were routine endoscopies (n = 268), while 40 emergency endoscopies were done within 6 h and 45 within 24 h of presentation.

**Table 1.**
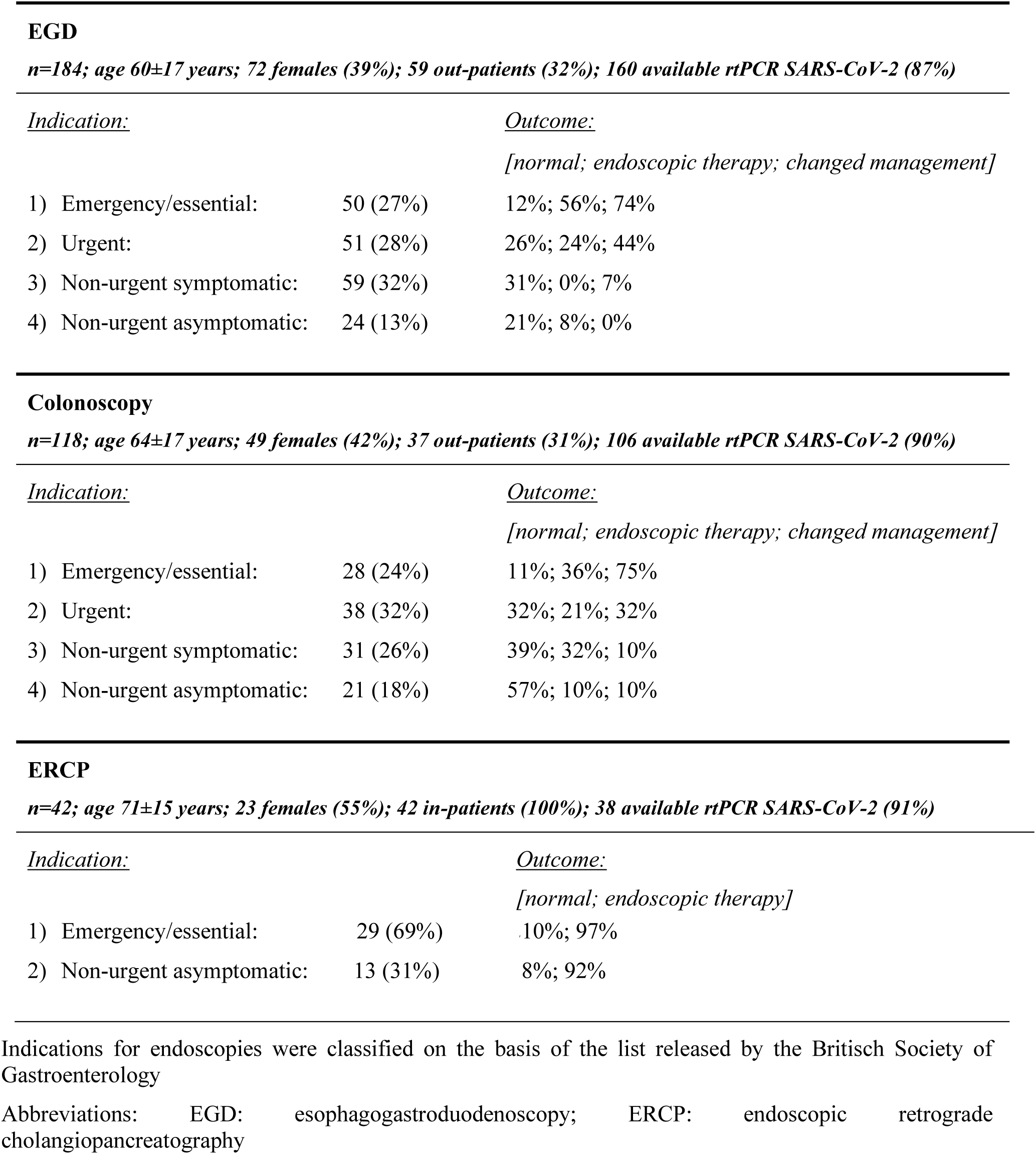
Endoscopic procedures performed between 23 March 2020 and 10 May 2020

| <u>Indication:</u> |  | <u>Outcome:</u> |
| --- | --- | --- |
|  |  | *[normal; endoscopic therapy; changed management]* |
| 1) Emergency/essential: | 50 (27%) | 12%; 56%; 74% |
| 2) Urgent: | 51 (28%) | 26%; 24%; 44% |
| 3) Non-urgent symptomatic: | 59 (32%) | 31%; 0%; 7% |
| 4) Non-urgent asymptomatic: | 24 (13%) | 21%; 8%; 0% |

**Colonoscopy***n=118; age 64±17 years; 49 females (42%); 37 out-patients (31%); 106 available rtPCR SARS-CoV-2 (90%)*
| <u>Indication:</u> |  | <u>Outcome:</u> |
| --- | --- | --- |
|  |  | *[normal; endoscopic therapy; changed management]* |
| 1) Emergency/essential: | 28 (24%) | 11%; 36%; 75% |
| 2) Urgent: | 38 (32%) | 32%; 21%; 32% |
| 3) Non-urgent symptomatic: | 31 (26%) | 39%; 32%; 10% |
| 4) Non-urgent asymptomatic: | 21 (18%) | 57%; 10%; 10% |

**ERCP***n=42; age 71±15 years; 23 females (55%); 42 in-patients (100%); 38 available rtPCR SARS-CoV-2 (91%)*
| <u>Indication:</u> |  | <u>Outcome:</u> |
| --- | --- | --- |
|  |  | *[normal; endoscopic therapy]* |
| 1) Emergency/essential: | 29 (69%) | 10%; 97% |
| 2) Non-urgent asymptomatic: | 13 (31%) | 8%; 92% |

Indications for endoscopies were classified on the basis of the list released by the British Society of Gastroenterology
Abbreviations: EGD: esophagogastroduodenoscopy; ERCP: endoscopic retrograde cholangiopancreatography

Performed interventions were categorized according to guidance published by the British Society of Gastroenterology (non-urgent procedures were subdivided into symptomatic and asymptomatic patients) [https://www.bsg.org.uk/covid-19-advice/endoscopy-activity-and-covid-19-bsg-and-jag-guidance/]. Overall, 200 procedures (56.6%) were classified as emergent, essential or urgent. Among emergent, essential or urgent upper endoscopies, 39.6% were therapeutic and 58.4% altered management (2.4% and 4.8%, respectively, for non-urgent procedures). For lower endoscopies, 27.3% of essential/emergent and urgent procedures were therapeutic and 50% altered management (23.1% and 9.6%, respectively, for non-urgent procedures). Five malignant tumours (frequency 1.4%) were diagnosed by upper (essential/urgent: 3; non-urgent: 2) or lower endoscopy (essential/urgent: 5; non-urgent: 0). Twelve non-urgent ERCPs were done to exchange biliary stents after maximal extension of exchange intervals, and all but two ERC(P)s were therapeutic.

The results of SARS-CoV-2 rtPCR were available prior to endoscopy in 313 procedures (88.6%). After implementation of structured testing, only eight from 302 (2.6%) non-emergency procedures were performed without known COVID-19 status. No patient scheduled for endoscopy tested positive for SARS-CoV-2 either before or after endoscopy. There occurred no infection of any endoscopy team member during the described period. Table 2 presents endoscopies performed in patients with and without rtPCR test results. A significantly higher proportion of patients without test results were investigated within 6 h of presentation (P < 0.001). However, there was no higher probability for a clinically significant finding or need for endoscopic haemostasis for emergency patients without test results (both P > 0.05).

**Table 2.** Comparison of endoscopic procedures in patients with or without known COVID-19 status

|  | **SARS-CoV-2 rtPCR**<br>**result available** | **SARS-CoV-2 rtPCR**<br>**result not available** |
| --- | --- | --- |
| **Patients (n)** | 313 (89%) | 39 (11%) |
| **Gender (m / f)** | 179 (57%) / 134 (43%) | 26 (67%) / 13 (33%) |
| **Age** | 62±18 (1 - 95) | 64±15 (30 - 91) |
| **In-patient / out-patient** | 231 (74%) / 82 (26%) | 25 (64%) / 14 (36%) |
| **Timing of endoscopy:** |  |  |
| *< 6h of symptom onset:* | 26 (8%) | 14 (36%) |
| *6-24h of symptom onset:* | 42 (13%) | 3 (8%) |
| *Routine:* | 245 (78%) | 22 (56%) |
| **Endoscopic Procedure:** |  |  |
| *EGD:* | 161 (51%) | 23 (59%) |
| *Colonoscopy:* | 106 (34%) | 12 (31%) |
| *ERCP:* | 38 (12%) | 4 (10%) |
| *EUS:* | 8 (3%) | 0 (0%) |
| **Outcome emergency procedures:** | **N=68** | **N=16** |
| *Clinically significant finding:* | 56 (82%) | 12 (75%) |
| *Normal finding:* | 9 (13%) | 1 (6%) |
| *Endoscopic haemostasis:* | 22 (32%) | 7 (44%) |
| *Biliary/pancreatic intervention:* | 13 (19%) | 3 (19%) |
| *Other endoscopic therapy:* | 3 (4%) | 1 (6%) |
| **Outcome routine procedures:** | **N=245** | **N=23** |
| *Clinically significant finding:* | 60 (25%) | 4 (17%) |
| *Normal finding:* | 68 (28%) | 13 (57%) |
| *Endoscopic haemostasis:* | 3 (12%) | 0 (0%) |
| *Biliary/pancreatic intervention:* | 28 (11%) | 1 (4%) |
| *PEG/feeding tube:* | 8 (3%) | 2 (9%) |
| *Polypectomy:* | 14 (6%) | 2 (9%) |
| *Other endoscopic therapy:* | 7 (3%) | 0 (0%) |

Clinically significant findings: acute life threatening conditions; result that changes patient management; gastrointestinal ulcers; polyps ≥10mm; reflux esophagitis ≥ Los Angeles C
Abbreviations: EGD: esophagogastroduodenoscopy; ERCP: endoscopic retrograde cholangiopancreatography EUS: endoscopic ultrasound; PEG: percutaneous endoscopic gastrostomy

## DISCUSSION

The reduction of endoscopies performed during COVID-19 pandemic is thought to (i) safe resources needed for COVID-19 patients, and (ii) minimize the risk of infection for patients and endoscopy team members. However, lock-down of endoscopy units during pandemic might confront us with potentially unmanageable numbers of patients afterwards. This strategy also implies that the pandemic of COVID-19 will soon be over, and this assumption might not necessarily be correct. Thus, we are in-need of simple and reliable screening tools that will allow functioning of endoscopy units without posing risks to patients or to the endoscopy teams. Overall, gastrointestinal endoscopy appears to be relatively safe when adequate protective measures are used (4, 5), but these measures significantly alter the workflow and potentially impair the quality of endoscopies.

Here we show that a moderate reduction of the endoscopy volume based on the structured rtPCR-based SARS-CoV-2 testing on nasopharyngeal swabs is feasible. The PECo strategy seems to work well, at least in regions with low to intermediate frequency of COVID-19 infections. However, the retrospective design, the lack of a structured follow-up for COVID-19 after the endoscopic procedure limit validity of the study. The reduction of endoscopy volume by 43% is substantially lower than the reported reductions of about 80% (1). Most endoscopies without a test result were performed in the evening or on weekends when results of routine tests were not available until the next morning. Based on our data and recent publications on the timing of endoscopy for upper gastrointestinal bleeding (6), the majority of endoscopies (except for suspected variceal haemorrhage with hemodynamic instability and severe septic shock due to cholangitis) can be safely deferred until the rtPCR result is available. Since false-negative rtPCR results are possible (7), we additionally recommend the use of standardized questionnaires to assess patients for COVID-19 symptoms and to adapt PPE. Our results might help to develop new strategies during future waves of COVID-19 infection or local outbreaks.

## Data Availability

The datasets used and analysed during the current study are available from the corresponding author on reasonable request.

## DECLARATIONS

### Ethics approval

This study was approved by the ethics committee of Ärztekammer des Saarlandes (Saarbrücken, Germany; #254/20). A waiver was obtained for the consent to participate in the study. The study was performed in accordance with the Declaration of Helsinki.

### Consent for publication

Not applicable

### Authors’ contributions

Markus Casper, Matthias Christian Reichert, Marcin Krawczyk: manuscript preparation and patient treatment

Jürgen Rissland, Sigrun Smola: virological diagnosis and manuscript preparation

Frank Lammert: manuscript revision, design of strategy and supervision of care

## ABBREVIATIONS

rtPCR: Reverse transcriptase polymerase chain reaction
ERCP: Endoscopic retrograde cholangiopancreatography
EUS: Endoscopic ultrasound
PECo: Pre-endoscopy SARS-CoV-2 testing
PEG: Percutaneous endoscopic gastrostomy
PPE: Personal protective equipment

